# Machine Learning for Interpretation of DNA Variants of Maturity-Onset Diabetes of the Young Genes Based on ACMG Criteria

**DOI:** 10.1101/2020.05.20.20108035

**Authors:** Yichuan Liu, Huiqi Qu, Adam S. Wenocur, Jingchun Qu, Xiao Chang, Joseph Glessner, Patrick Sleiman, Lifeng Tian, Hakon Hakonarson

**Affiliations:** Center for Applied Genomics, Children’s Hospital of Philadelphia, Philadelphia, PA, USA; Department of Human Genetics, Children’s Hospital of Philadelphia, Philadelphia, PA, USA; Division of Human Genetics, Department of Pediatrics, The Perelman School of Medicine, University of Pennsylvania, Philadelphia, PA, USA

## Abstract

**Background:** Maturity-onset diabetes of the young (MODY) is a group of dominantly inherited monogenic diabetes, with *HNF4A-MODY*, *GCK-MODY* and *HNF1A-MODY* being the three most common genes responsible. Molecular diagnosis of MODY is important for precise treatment. While a DNA variant causing MODY can be assessed by the criteria of the American College of Medical Genetics and Genomics (ACMG) guidelines, gene-specific assessment of disease-causing mutations is important to differentiate between the MODY subtypes. As the ACMG criteria were not originally designed for machine learning algorithms, they are not true independent variables.

**Methods:** In this study, we applied machine learning models for interpretation of DNA variants in MODY genes defined by the ACMG criteria based on Human Gene Mutation Database (HGMD) and ClinVar.

**Results:** The results show highly predictive abilities with accuracy over 95%, suggest that this model could serve as a fast, gene-specific method for physicians or genetic counselors assisting with diagnosis and reporting, especially when confronted by contradictory ACMG criteria. Also, the weight of the ACMG criteria shows gene specificity which advocates for the application of machine learning methods with the ACMG criteria to capture the most relevant information for each disease-related variant.

**Conclusion:** Our results highlight the need for different weights of the ACMG criteria in relation with different MODY genes for accurate functional classification. For proof of principle, we applied the ACMG criteria as feature vectors in a machine learning model obtaining precision-based result.

## Background

Monogenic diabetes results from DNA mutations in a single gene and accounts for about 1 to 4 percent of all cases of diabetes in United States (https://www.niddk.nih.gov/health-information/diabetes/overview/what-is-diabetes/monogenic-neonatal-mellitus-mody). The most common form of monogenic diabetes is maturity-onset diabetes of the young (MODY), an autosomal dominant disease that most commonly occurs in adolescence or early adulthood[1]. Genetic sequencing is needed to identify the casual mutations and diagnose different types of MODYs[2]. The DNA variant causing MODY can be specifically assessed by the criteria established by the American College of Medical Genetics and Genomics (ACMG), as published in their guidelines[3]. While the ACMG guidelines can be universally applied for all human DNA variants, our previous study suggests that a gene-specific assessment is important for disease-causing mutations in different MODY genes[4]. In addition, contradictory evidence is commonly seen in functional classification of genetic variation by the ACMG guidelines[5]. While the ACMG guidelines may suggest a variant of uncertain significance (VUS), classification of the variant may have contradictory evidence, and some variants with contradictory evidence may turn out to have reliable definite classification.

In this study, we focused on DNA variants of three MODY genes (i.e., *HNF1A, HNF4A*, and *GCK)* underlying the three most common types of MODYs[6], where we tested machine learning models for interpretation of DNA variants, using the ACMG criteria. Our results highlight the need for a different weight of the ACMG criteria in the functional classification of DNA variants of different MODY genes.

## Methods

### Data Collection for Machine Learning Procedures

Known DNA variants of the three MODY genes, *HNF1A, HNF4A*, and *GCK*, were acquired from the database of common SNP 151[7], the ClinVar database[8], and the Human Gene Mutation Database (HGMD) 2019 professional version[9]. Among the multi-hundred variants reported in these genes, approximately one half have the classification of pathogenic/likely pathogenic(P/LP) variants according to the annotation in ClinVar or HGMD. According to the HGMD, the three genes were curated by Professor Andrew Hattersley, a leading genetic expert in MODYs (http://www.hgmd.cf.ac.uk/docs/newback.html). The classification of benign/likely benign (B/LB) variants varies between the different databases according to the annotation of ClinVar or common SNP 151. Overall, for the three genes, there are 899 unique variants reported in *HNF1A*, including 569 P/LP sites, and 330 B/LB sites; 1037 unique variants for *HNF4A*, including 182 P/LP sites, and 855 B/LB sites; and 1664 unique variants for *GCK*, including 1065 P/LP sites and 599 B/LB sites. However, a number of these variants have different annotation features between the different databases.

### Feature Vector Generation

The feature vectors for the machine learning modeling are the criteria based on the ACMG guidelines[10]. The criteria terms were generated based on InterVar[11], a computational tool that uses a pre-annotated or VCF file as an input and generates automated interpretation based on the ACMG criteria. It should be noted that not all 33 ACMG criteria can be computationally scored. For example, the PS3 criteria requires well-established *in vitro* or *in vivo* functional studies supportive of a damaging effect on the gene or gene product. As a result, the number of ACMG criteria, or the length of feature vectors for the three MODY genes is 15, include PVS1, PS1, PS4, PM1, PM2, PM4, PM5, PP2, PP3, PP5, BA1, BS1, BP4, BP6, and BP7.

Using machine learning regression procedures, we normalized the weights for the evidence of different categories in accordance with the ACMG guidelines, i.e., assuming that the weight coefficient of PVS1 criteria is 1, PS criteria is 1/2, PM criteria is 1/6 and PP criteria is 1/12. We additionally assumed that the weight coefficient of BA1 is −1, BS criteria is −1/2 and BP criteria is −1/4. As the ACMG criteria were not originally designed for machine learning, these criteria are not true independent variables. Multi-collinearity among feature vectors is commonly seen within each gene and an example is the PM1 and PP2 criteria. By definition, a PM1 hit means the variant locates in a mutational hot spot and/or critical and well-established functional domain without benign variation; PP2 hit means there is a missense variant in a gene that has a low rate of benign missense variation and in which missense variants are a common mechanism of disease. In many situations PM1 and PP2 are consistent with each other, which has the potential to cause mis-weighing of the two criteria because of multi-collinearity. In order to detect the collinearity among feature vectors, we calculated the variance inflation factor (VIF) and pairwise correlation coefficient for the ACMG criteria. Feature vectors with VIF greater than 10 or correlation coefficient larger than 0.8 were removed before the learning procedures.

### Learning Procedures & Predictive Modeling

The machine learning procedure is a typical logistic regression based on the Scikit-learn package in Python[12]. For the detection of weight matrix of the ACMG criteria, all the variants including P/LP and B/LB were taken into account. For predictive modeling, we split the data based on 2fold random shuffle processes. In other words, the P/LP and B/LB variants were split randomly into equal size sets, with one set serving as training data, and the other set serving as testing data to determine the predictive capabilities of the model. This process was repeated ~20 times to obtain the mean and standard deviation for accuracy measures, including sensitivity and specificity.

## Results

### Weight matrix of the ACMG criteria show differences among the three MODY genes

Based on the machine learning procedure, we found that the weight matrix of the ACMG criteria was significantly different between the three MODY genes, *HNF1A, HNF4A* and *GCK* (Table 1, Figure 1). The differences are non-trivial and must be taken into consideration in clinical interpretation of DNA variants for genetic diagnosis.

**Figure 1.**
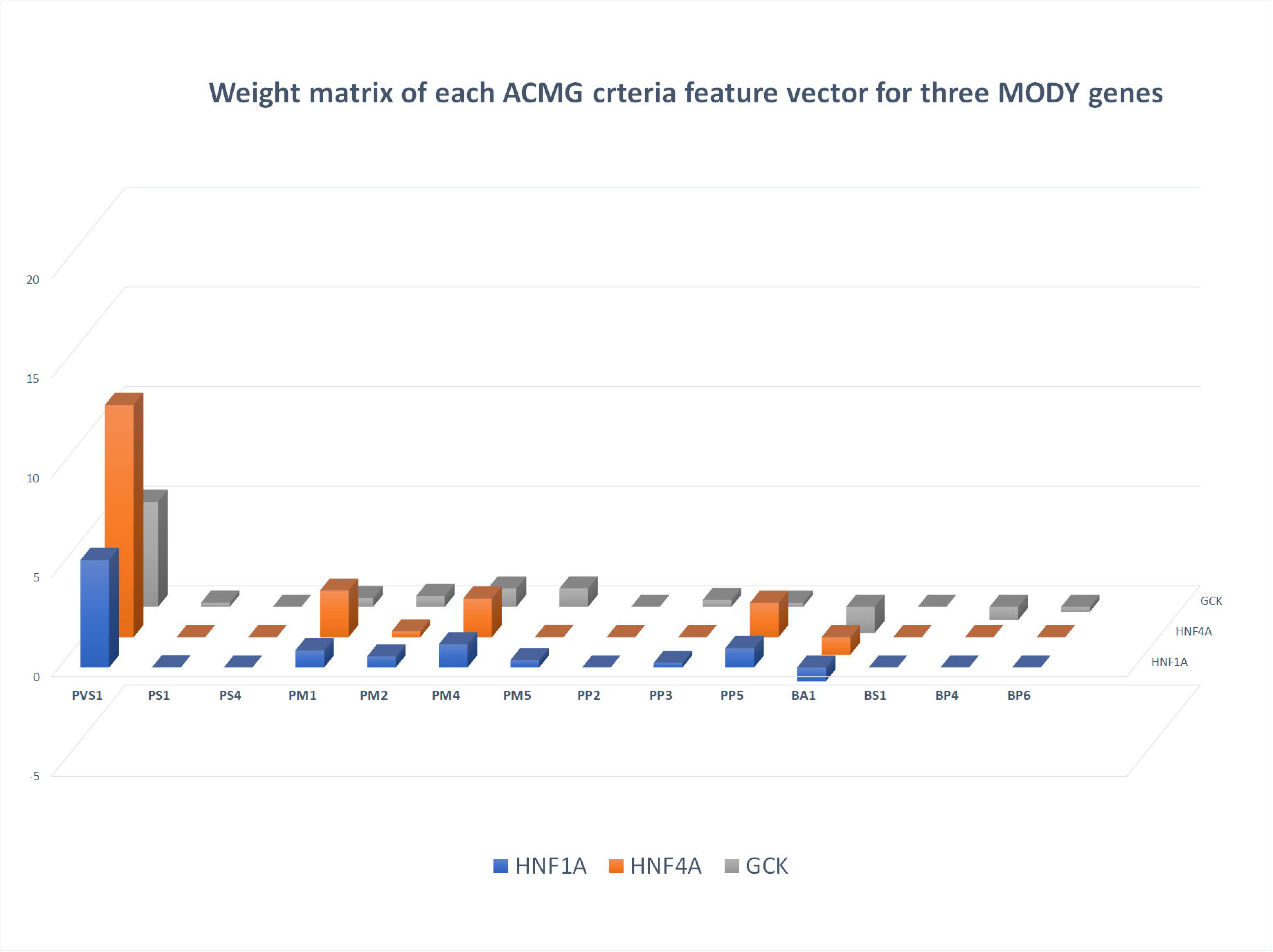
Weight matrix of three MODY genes, HNF1A, HNF4A and GCK: normalized weight for ACMG criteria for three most common MODY genes.

**Table 1.** Weight matrix of the ACMG criteria for three MODY genes

| Term | Frequency |  |  |  |  |  | weight |  |  |
| --- | --- | --- | --- | --- | --- | --- | --- | --- | --- |
|  | HNF1A<br>PLP | HNF1A<br>BLB | HNF4A<br>PLP | HNF4A<br>BLB | GCK<br>PLP | GCK<br>BLB | HNF1A | HNF4A | GCK |
| **PVS1** | 0.3040 | 0.0030 | 0.2582 | 0.0000 | 0.2183 | 0.0033 | 5.401 | 11.665 | 5.275 |
| **PS1** | 0.0018 | 0.0000 | 0.0000 | 0.0000 | 0.0039 | 0.0000 | 0.018 | 0.000 | 0.210 |
| **PS4** | 0.0000 | 0.0030 | 0.0000 | 0.0000 | 0.0000 | 0.0000 | 0.000 | 0.000 | 0.000 |
| **PM1** | 0.5149 | 0.0030 | 0.4835 | 0.0000 | 0.6326 | 0.0083 | 0.866 | 2.340 | 0.442 |
| **PM2** | 0.9772 | 0.4273 | 0.7802 | 0.1450 | 0.9922 | 0.5092 | 0.555 | 0.290 | 0.542 |
| **PM4** | 0.0053 | 0.0000 | 0.0055 | 0.0000 | 0.0049 | 0.0000 | 1.167 | 1.944 | 0.923 |
| **PM5** | 0.0264 | 0.0000 | 0.0055 | 0.0000 | 0.1335 | 0.0000 | 0.375 | 0.007 | 0.927 |
| **PP2** | 0.0000 | 0.0000 | 0.5440 | 0.0012 | 0.0000 | 0.0000 | 0.000 | 0.000 | 0.000 |
| **PP3** | 0.6151 | 0.0061 | 0.5769 | 0.0000 | 0.7437 | 0.0050 | 0.235 | 0.000 | 0.347 |
| **PP5** | 0.1195 | 0.0000 | 0.1923 | 0.0000 | 0.1540 | 0.0033 | 0.975 | 1.727 | 0.202 |
| **BA1** | 0.0105 | 0.2091 | 0.0165 | 0.1708 | 0.0039 | 0.1703 | -0.696 | -0.880 | -1.312 |
| **BS1** | 0.0228 | 0.5667 | 0.0440 | 0.5895 | 0.0078 | 0.4891 | 0.000 | 0.000 | 0.000 |
| **BP4** | 0.0193 | 0.0727 | 0.0165 | 0.0222 | 0.0458 | 0.0234 | 0.000 | 0.000 | -0.665 |
| **BP6** | 0.0123 | 0.1182 | 0.0220 | 0.0480 | 0.0029 | 0.0234 | 0.000 | 0.000 | -0.253 |
| **BP7** | 0.0123 | 0.0636 | 0.0110 | 0.0211 | 0.0400 | 0.0200 | 0.000 | 0.000 | 0.000 |

PS evidence are rarely observed with MODY variants. On the other hand, PS4 evidence (the prevalence of the variant in affected individuals is significantly increased compared with the prevalence in controls) is commonly observed and often misclassified. As an example, the *HNF1A* variant, 12:121420807-G-A, or rs1183910, has been reported in a genome-wide association study (GWAS) to be associated with C-reactive protein, a marker of inflammation. [13]. However, as a common SNP with a minor allele frequency of 0.292 in European populations, it can’t be a variant causing the rare and dominantly inherited form of *HNF1A*-MODY.

Among the PM evidence, PM1, which is defined as located in a mutational hot spot and/or in a critical and well-established functional domain (e.g., active site of an enzyme) without benign variation, and PM2 (absent from controls or at extremely low frequency if recessive in Exome Sequencing Project, 1000 Genomes Project, or Exome Aggregation Consortium) are both commonly observed in support of pathogenic variants in the three MODY genes. However, PM2 is also commonly seen in B/LB variants in these three genes, thus lacking specificity for the functional classification. In our study, PM2 showed a VIF of 79.0 in *HNF1A*, and 247 in *GCK*. As a result, although PM2 is much more common than PM1 in the three MODY genes, the weight of PM2 in *HNF1A* is lower than PM1.

Among the PP evidence, PP2 (missense variant in a gene that has a low rate of benign missense variation and in which missense variants are a common mechanism of disease) is absent in *HNF1A* and *GCK*, but commonly seen in *HNF4A*. However, PP2 has a correlation coefficient of 0.932 with PM1, so it doesn’t add much weight to the classification of P/LP variants in *HNF4A*.

### Highly accurate predictive ability for MODY gene pathogenicity

*HNF4A*-MODY (MODY1), *GCK*-MODY (MODY2), and *HNF1A*-MODY (MODY3) are the three most common types of MODYs accounting for ~70% of all MODY genes[14]. Therefore, predictive model aimed at the recognition of pathogenic variants would be useful for the diagnosis of novel mutations in these genes. As described in the method section, we used 2-fold random shuffle testing which used 50% of the mutations as training data, another half as testing data, and repeated the analysis dozen times. The logistic regression machine learning shows overall accuracy above 95% for MODY gene mutations (Figure 2). Both *HNF1A* and *HNF4A* have specificity close to 100%, and specificity in *GCK* is also above 95%. The lower specificity however is also consistent with the benign phenotype and mild clinical expression of *GCK-*MODY. The results prove the principle that ACMG criteria could be applied as meaningful feature vectors in machine learning model and machine learning model based on ACMG criteria could provide accurate pathogenic classification for other Mendelian disease genes in a gene-specific way.

**Figure 2.**
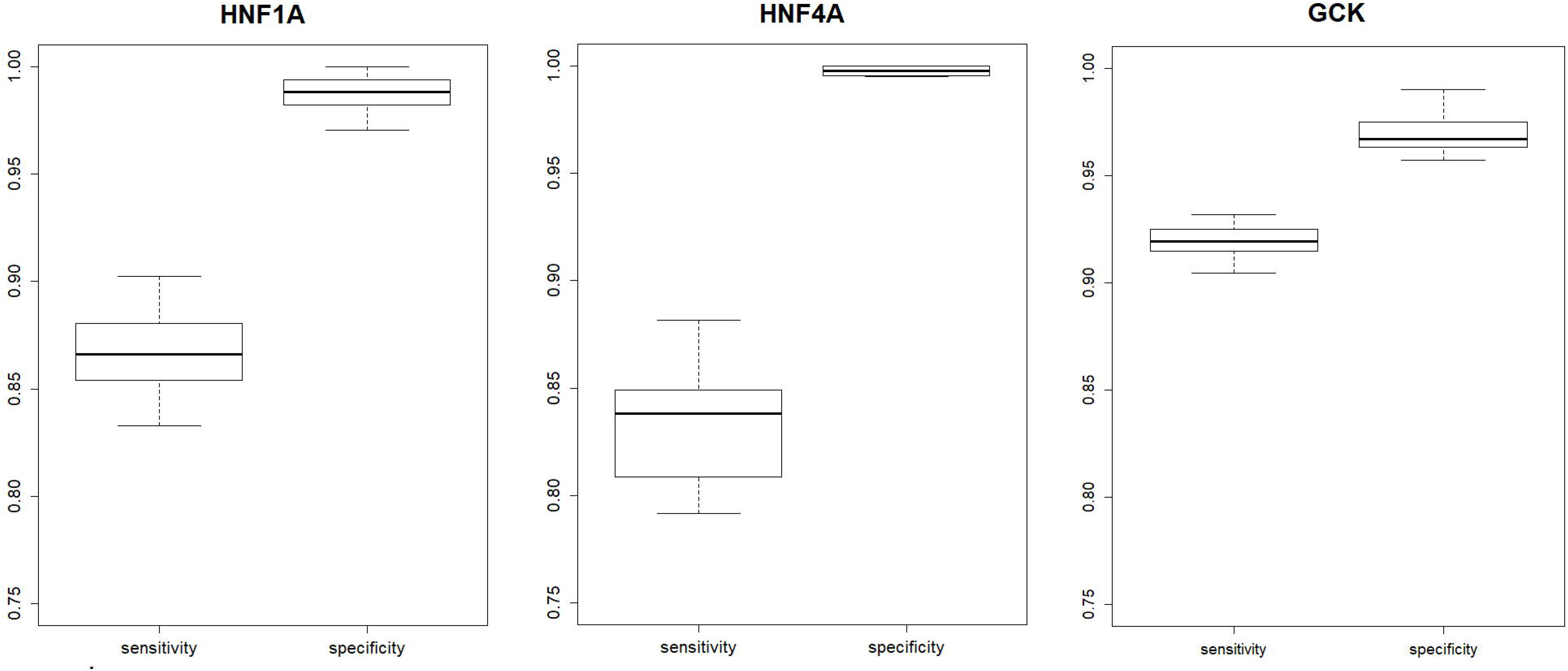
Overall accuracy based on logistic regression machine learning tests. Boxplot represents the sensitivity and specificity for 2-fold random shuffle tests.

## Discussion

In the past decade, sequencing technologies have evolved rapidly with the advance of high-throughput next-generation sequencing (NGS). By adopting NGS, clinical laboratories are now performing an ever-increasing volume of genetic testing for genetic disorders. Increased complexity in genetic testing has been accompanied by new challenges in sequence interpretation and multiple new standards have been implemented for physicians and genetic counselors regarding interpretation and reporting of sequence variants at different level of pathogenicity. Currently there are multiple computational tools which are based on different algorithms and databases that are being used to predict the pathogenicity of DNA variants, such as SIFT[15], MutationTaster[16], likelihood ratio test (LRT)[17], FATHMM by supervised machine learning model [18], GERP++ by maximum likelihood evolutionary rate estimation [19] for coding variants, and DANN for both coding and non-coding variants using deep neural network [20].

However, all these computational tools assess each gene with a common rule which is not based on biology, while our study suggested gene-specific assessment for pathogenicity is required at least for MODY genes[4]. The evolutionary selection pressures on MODYs vary across different genes, whereas it is minimum in the case of GCK-MODY[21]. Similar issues exist with functional classification by the ACMG criteria which are globally applied for all human genes. The ACMG Criteria contain 33 terms that lead to five categories of mutations (“pathogenic,” “likely pathogenic,” “uncertain significance,” “likely benign,” and “benign”), is one of the most commonly used standards.

Maturity-onset diabetes of the young (MODY) is a group of dominantly inherited monogenic diabetes, and *HNF4A*-MODY (MODY1), GCK-MODY (MODY2), and *HNF1A-*MODY (MODY3) are the three most common types of MODY. These MODY genes are involved in different molecular pathways. MODY variants of different gene show different clinical features and require different treatment. For example, HNF1A-MODY has reduced beta cell mass or impaired function, and has been treated with sulfonylureas with excellent results for decades[22]. These patients are so sensitive to sulfonylurea treatment and may be susceptible to develop hypoglycemia during sulfonylurea treatment[22]. *HNF4A*-MODY has similar clinical features with HNF1A-MODY, and the affected transcription network plays a role in the early development of the pancreas. The pancreatic beta cells produce adequate insulin in infancy but capacity for insulin production declines thereafter[23]. The beta cells in GCK-MODY have a normal capacity to make and secrete insulin, but do so only above an abnormally high glucose threshold which produces a chronic, mild increase in blood sugar, which is usually asymptomatic[21]. The treatment of GCK-MODY can be achieved by a healthy diet and exercise, while oral hypoglycemic agents or insulin is of no benefit in these patients[21]. Therefore, molecular diagnosis of these MODYs are important for precise treatment.

## Conclusion

In this study, we applied a computational machine learning method together with the ACMG criteria for functional classification of genetic variants of the three most common MODY genes, *HNF1A, HNF4A* and *GCK*. Our results show that a typical machine learning model using 15 computational ACMG criteria as the feature vector has predictive abilities that are highly accurate (>95% accuracy) for hundreds of annotated variants in three MODY genes. It suggests that this model could serve as a fast, gene-specific method for physicians or genetic counselors assisting with diagnosis and reporting, especially when confronted by contradictory ACMG criteria. Also, we show that the weight of the ACMG criteria shows gene specificity which advocates for the application of machine learning methods with the ACMG criteria to capture the most relevant information for each disease-related variant.

## Data Availability

All data referred to in the manuscript are publicly available from these databases:
HGMD 2019 version: http://www.hgmd.cf.ac.uk/ac/index.php
ClinVar: https://www.ncbi.nlm.nih.gov/clinvar/
Common SNP 151: https://www.ncbi.nlm.nih.gov/snp/

## Novelty statement

Machine learning based on ACMG criteria for all known MODY causal genetic variants show different ACMG weight for different MODY genes. ACMG criteria have high accuracy predictive abilities for functional MODY genetic variants. Our results highlight the need for different weights of the ACMG criteria in relation with different MODY genes for accurate functional classification. For proof of principle, we applied the ACMG criteria as feature vectors in a machine learning model obtaining precision-based result.

## Ethics approval and consent to participate

None

## Consent to publish

Not applicable

## Availability of data and materials

All data referred to in the manuscript are publicly available from these databases:

HGMD 2019 version: http://www.hgmd.cf.ac.uk/ac/index.php

ClinVar: https://www.ncbi.nlm.nih.gov/clinvar/

Common SNP 151: https://www.ncbi.nlm.nih.gov/snp/

## Authors’ Contributions

Dr. Yichuan Liu, and Dr. Huiqi Qu conceptualized and designed the study, drafted the initial manuscript, and reviewed and revised the manuscript. Dr. Adam S. Wenocur, Mr. Jingchun Qu, Dr. Xiao Chang, Dr. Joseph Glessner, Dr. Patrick Sleiman, and Dr. Lifeng Tian collected data, carried out the initial analyses, and reviewed and revised the manuscript. Dr. Hakon Hakonarson conceptualized and designed the study and critically reviewed the manuscript.

## Acknowledgement

We thank the Center for Applied Genomics (CAG) staff and supports from Children’s Hospital of Philadelphia (CHOP)

